# Platelet activation with the Aeson Bioprosthetic Total Artificial Heart: insight from aspirin treatment and outcomes

**DOI:** 10.1101/2023.01.20.23284588

**Authors:** David M. Smadja, Christophe Peronino, Léa Jilet, Peter Ivak, Yuri Pya, Aurélien Philippe, Christian Latremouille, Finn Gustafsson, Faiz Z. Ramjankhan, Jean Christian Roussel, André Vincentelli, Erwan Flecher, Pascale Gaussem, Piet Jansen, Ivan Netuka

## Abstract

**Introduction:** The Aeson bioprosthetic total artificial heart (A-TAH) is a pulsatile and autoregulated device. The aim of this study was to evaluate the level of platelet activation secondary to A-TAH implantation.

**Methods:** We examined the level of platelet activation markers in adult patients receiving A-TAH support (n=16) during clinical follow-up by quantifying sP-selectin (sP-sel) and sCD40L in plasma.

**Results:** The cumulative duration of A-TAH support was 3587 days. Before implantation, sCD40L was 2684.6 pg/mL (954.0-16706.1) and remained steady after implantation [3305.8 pg/mL (1234.2-12327.5), 3300.5 pg/mL (1041.5-8370.1), and 2560.0 pg/mL (1325.5-14039.5), respectively, at <3 months, 3–6 months and >6 months; non-significant difference along time-period]. sP-sel was 33997.0 pg/mL (16019.6-73377.6) and remained steady after implantation [33580.1 pg/mL (13979.8-53395.2), 33204.9 pg/mL (15332.6-67263.4), and 34684.5 pg/mL (14084.9-49206.0), respectively, at <3 months, 3–6 months and >6 months; non-significant difference along time-period]. Levels sP-sel and sCD40L analysed according to aspirin or heparin use did not change. Finally, no relation existed between pericardial effusion and aspirin use according to timing of aspirin start and drain removal.

**Conclusions:** We demonstrated that A-TAH does not induce significant platelet activation. Absence of relationship between aspirin and platelet activation levels or pericardial effusion may support potential for withdrawal of antiplatelet therapy after A-TAH implantation in the future.

## Introduction

Mechanical circulatory support (MCS) is now a mainstay in the treatment of advanced heart failure as bridge to transplantation or destination therapy. This therapeutic area has undergone many evolutions in recent years with the magnetically-levitated Heartmate 3 left ventricular assist device (HM3, Abbott Laboratories, Chicago, IL) ^12^ and the autoregulated bioprosthetic Aeson® total artificial heart (A-TAH, Carmat SA, Velizy-Villacoublay, France) ^3^. Historically, MCS was associated with numerous hemocompatibility-related adverse events including thrombosis, stroke and bleeding ^4^. Therefore, anti-thrombotic therapy is mandatory during MCS to decrease the risk of thromboembolic complications. For most long term MCS, a dual therapy is used consisting of an anti-platelet agent and a vitamin K antagonist (VKA) at therapeutic doses. In A-TAH we previously demonstrated that anti-thrombotic therapy may be satisfactorily provided with low molecular weight heparin (LMWH) and aspirin during the first 6 months ^3^. In the European A-TAH pivotal study, we described that anticoagulation with intermediate LMWH doses efficiently prevented coagulation activation ^5^. Similar to recommended protocols for left ventricular assist devices, the guideline for Aeson implantation suggests starting aspirin four days after chest drain removal.

In LVAD recipients, the risk of bleeding resulting from an acquired von Willebrand syndrome affecting primary hemostasis has prompted several centers, in particular in Europe, to decrease the dose or to withdraw antiplatelet agents ^6^. The European TRACE study ^7^ proposed a VKA monotherapy to reduce the incidence of major bleeding without increasing thrombotic events during Heartmate II (HM2) implantation. These results have been confirmed in the PREVENTII trial ^8^. Moreover, the MOMENTUM 3 trial demonstrated that usual and low aspirin doses had same rates of bleeding and thrombotic events after HM3 implantation ^9^ and some reports illustrate the absence of thrombotic events in patients without antiplatelet therapy ^10,11^. In patients supported by A-TAH, we previously described a decrease in platelet activation evaluated with platelet microvesicles in three patients implanted with A-TAH compared to a HM2 cohort ^12^.

The aim of our study was to evaluate safety of aspirin after A-TAH implantation pas well as platelet activation evaluated by levels of soluble P-selectin (sP-sel) and CD40 ligand (sCD40L) before A-TAH implantation and during the whole follow-up after implantation.

## Patients and methods

### Patients and samples

Sixteen patients implanted with A-TAH were included in this study (**Table 1** for patient characteristics) and provided informed consent (*<u>NCT02962973;</u>* Ethics committee/IRB Ile de France III gave ethical approval for this study: IRB 2016-A00504-47). Blood samples were collected on EDTA once a week during ICU, once a month during the first 6 months after implantation and every three months thereafter. Platelet poor plasma (PPP) was obtained after centrifugation at 2500*g* for 15 minutes and stored at − 80 °C until analysis. Anti-thrombotic treatment started with un-fractionated heparin I.V. (UFH) post-implant after chest closure and when chest tube drainage was below 50 ml/h mL for four consecutive hours. UFH efficacy in the therapeutic range of 0.2 – 0.3 IU/mL was monitored by Anti-Xa activity. Acetylsalicylic acid (Aspirin 75 or 100 mg) was added 4 days after chest drain removal pending absence of active bleeding. When renal function was normalized (creatinine clearance > 30 ml/min; Cockcroft & Gault formula), and if there was no indication for imminent invasive procedures, UFH was switched to LMWH (tinzaparin 90-175 IU/kg/24h or enoxaparin 75-150 IU/Kg/24h). All data produced in the present study are available upon reasonable request to the authors.

**Table 1:** Patients description.

| Patients | 1 | 2 | 3 | 4 | 5 | 6 | 7 | 8 | 9 | 10 | 11 | 12 | 13 | 14 | 15 | 16 |
| --- | --- | --- | --- | --- | --- | --- | --- | --- | --- | --- | --- | --- | --- | --- | --- | --- |
| Gender | Male | Male | Male | Male | Male | Male | Male | Male | Male | Male | Male | Male | Male | Male | Male | Male |
| BMI kg/m<sup>2</sup> | 24.69 | 27.04 | 30.08 | 25.43 | 26.7 | 29.3 | 22.04 | 28.73 | 27.14 | 30.13 | 24.16 | 26.33 | 25.65 | 21.91 | 23.35 | 23.89 |
| BSA (m<sup>2</sup>) | 2 | 1.96 | 2.06 | 1.93 | 2.13 | 2.2 | 1.89 | 2 | 2.06 | 2.36 | 1.9 | 1.91 | 1.77 | 1.88 | 1.86 | 2.02 |
| Cardiac index (L/min/m<sup>2</sup>) | 2 | 2.15 | 1.55 | 1.76 | 1.24 | 1.34 | 1.66 | 1.74 | 1.6 | 1.65 | 1.54 | 1.78 | 1.8 | 2.6 | 4.08 | 2.19 |
| Intermacs Class | 3. Stable but inotrope dep. | 3. Stable but inotrope dep. | 3. Stable but inotrope dep. | 3. Stable but inotrope dep. | 4. Resting symptoms | 3. Stable but inotrope dep. | 3. Stable but inotrope dep. | 2. Progressive decline | 3. Stable but inotrope dep. | 3. Stable but inotrope dep. | 3. Stable but inotrope dep. | 3. Stable but inotrope dep. | 3. Stable but inotrope dep. | 3. Stable but inotrope dep. | 3. Stable but inotrope dep. | 4. Resting symptoms |
| Indication BTT ou DT | DT | BTT | BTT | DT | DT | BTT | BTT | BTT | DT | BTT | DT | DT | BTT | BTT | BTT | DT |
| Support Duration (days) | 47 | 243 | 75 | 270 | 772 | 155 | 304 | 109 | 271 | 308 | 421 | 36 | 121 | 96 | 128 | 231 |
| preoperative creat (mg/dL) | 2.69 | 1.24 | 1.64 | 0.88 | 1.39 | 1.23 | 0.8 | 1.04 | 1.13 | 1.14 | 1.31 | 2.05 | 1.57 | 0.8 | 1.4 | 1.91 |
| heart failure cause |  |  |  |  |  |  |  |  |  |  |  |  |  |  |  |  |
| dilated | Yes | Yes | No | Yes | Yes | No | No | No | No | Yes | Yes | No | Yes | Yes | No | No |
| ischemic | No | No | Chronic | No | No | No | Chronic | Acute | Chronic | No | No | Chronic | No | No | Chronic | No |
| other | No | No | No | No | No | Dilated (Family type) | No | No | No | No | No | No | No | No | No | Giant Cell Myocarditis |
| comorbidities |  |  |  |  |  |  |  |  |  |  |  |  |  |  |  |  |
| arterial hypertension | No | No | No | No | No | No | No | Yes | No | Yes | Yes | No | No | No | No | No |
| diabetes | No | No | No | No | No | ND | No | No | Yes | Yes | No | Yes | Yes | No | Yes | No |
| dyslipidemia | No | No | No | No | No | No | Yes | Yes | No | No | No | No | No | No | Yes | No |
| **Hemostatic complication after implantation** |  |  |  |  |  |  |  |  |  |  |  |  |  |  |  |  |
| thrombosis | No | No | No | No | No | No | No | No | No | No | No | No | No | No | No | No |
| bleeding (excluding perioperative bleeding) | No | No | No | No | No | No | No | Yes | No | No | Yes | No | Yes | Yes | Yes | Yes |
| pericardial effusion | No | Yes | No | Yes | No | No | Yes | No | No | Yes | Yes | No | Yes | Yes | No | No |

*Dep.: dependent; BTT: bridge to transplantation; DT: Destination therapy*

### Biomarkers of platelet activation

Soluble P-selectin (sP-sel) and soluble CD40 ligand (sCD40L) levels were quantified in platelet-poor plasma (PPP) using a Human Magnetic Luminex Assay (R&D Systems, Minneapolis, MN). Data were assessed with the Bio-Plex 200 using the Bio-Plex Manager 5.0 software (Bio-Rad, Marnes-la-Coquette, France).

### Statistical analyses

For descriptive analysis, categorical data were expressed using counts and percentages, and continuous data using either means and standard errors or medians, interquartile ranges, and ranges, as appropriate. Evaluation of changes over time of biomarker levels (CD40, P-Selectin) was tested using linear mixed effect models including the value of biomarkers as dependent variable, the period of time as explanatory variables and the patient as random effect. We tested the association between the levels of biomarkers and treatments (aspirin or heparin) using also linear mixed effect models including the value of biomarkers as dependent variable, the treatment as explanatory variable and the patient as random effect. A Fisher’s exact test was used to compare the occurrence of pericardial effusion according to aspirin starting point (before or after drain removal) and a Mann-Whitney test was used to compare the number of re-interventions for pericardial effusion. All tests were two-sided with significance level set atm 0.05. Statistical analyses were performed using SAS Enterprise Guide version 8.3 (© 2019-2020, SAS Institute Inc., Cary, NC, USA).

## Results

### A-TAH implantation did not induce platelet activation

Sixteen male patients of mean age of 58.15 years (SD=12.1) were implanted with the A-TAH between September 2016 and November 2021. The indication was bridge to transplantation (BTT) for 9 out of 16 patients (56.25%) and destination therapy (DT) for 7/16 patients (43.75%); all patients were classified as INTERMACS Profiles 2 to 4, with a mean cardiac index 1.91 (SD= 0.66) liters/min/m^2^. The median follow-up duration was 224 (range 36 to 772) days (**Table 1**). No arterial thrombotic events were reported. One patient experienced a deep vein thrombosis (DVTs) attributed to a pre-implant central venous catheter. The cumulative support duration was 3587 days. We compared platelet activation biomarkers sP-sel and sCD40L in a total of 174 samples collected before and after implantation and during the follow-up as previously described ^13^. Platelet counts did not significantly change over time and did not differ at <3 months, 3–6 months and >6 months after implantation compared to reference values before A-TAH (**Table 2 and Figure 1**). Before implantation, sCD40L was 2684.6 pg/mL (954.0-16706.1) and remained steady after implantation (**Table 2:** 3305.8 pg/mL (1234.2-12327.5), 3300.5 pg/mL (1041.5-8370.1), and 2560.0 pg/mL (1325.5-14039.5), respectively,at <3 months, 3–6 months and >6 months; non-significant difference along time-period). sP-sel was 33997.0 pg/mL (16019.6-73377.6) and remained steady after implantation (**Table 2:** 33580.1 pg/mL (13979.8-53395.2), 33204.9 pg/mL (15332.6-67263.4), and 34684.5 pg/mL (14084.9-49206.0), respectively, at <3 months, 3–6 months and >6 months; non-significant difference along time-period). Platelet counts and levels of sCD40L and sP-sel are illustrated in **Figure 1**. Since we previously demonstrated the relevance of LMWH or heparin as optimal treatment after A-TAH implantation ^3,5^ to avoid any disseminated intravascular coagulation and associated thrombocytopenia we also compared sP-sel and sCD40L levels according to the presence or the absence of anticoagulation. As shown in Table 3, LMWH/heparin did not influence biomarker levels.

**Table 2:** Absence of modification in platelet activation biomarkers and platelet count after Aeson bioprosthetic total artificial heart (Aeson; A-TAH) implantation in 16 patients. Values as median (range). The results of the linear mixed model are expressed as estimate (standard error) and p-value.

|  | *Before implant* |  | *< 3 months* |  | *3 – 6 months* |  | *6 – 10 months* |  |
| --- | --- | --- | --- | --- | --- | --- | --- | --- |
|  | *n=16 (1 sample /patient)* |  | *n=99 (6.2 samples/patient [1-9])* |  | *n=44 (2.8 samples/patient [0-7])* |  | *n=15 (0.8 sample/patient [0-7])* |  |
| **Platelets (10<sup>9</sup>/L)** | 190 (130-671.0) | Ref | 238.0 (116.0-525.0) | -44.42 (17.08),<br>p-value=0.0104 | 212.0 (116.0-540.0) | -23.54 (19.62), p-value=0.2324 | 199.5 (95.0-496.0) | -27.64 (26.39),<br>p-value=0.2969 |
| **CD40 levels (pg/mL)** | 2684.6 (954.0-16706.1) | Ref | 3305.8 (1234.2-12327.5) | 77.89 (505.36),<br>p-value=0.8777 | 3300.5 (1041.5-8370.1) | 453.81 (556.36),<br>p-value=0.4159 | 2560.0 (1325.5-14039.5) | 264.56 (708.63),<br>p-value=0.7094 |
| **P-Selectin levels (pg/mL)** | 33997.0 (16019.6-73377.6) | Ref | 33580.1 (13979.8-53395.2) | 2691.01 (2089.27), p-value=0.1996 | 33204.9 (15332.6-67263.4) | 3226.73 (2299.46),<br>p-value=0.1624 | 34684.5 (14084.9-49206.0) | 4550.92 (2927.36),<br>p-value=0.1220 |

**Table 3:** sP-sel and sCD40L are not influenced by anticoagulation therapy after Aeson bioprosthetic total artificial heart (Aeson; A-TAH) implantation in 16 patients. *p-values from the linear mixed model

|  | Medication |  |  |  |
| --- | --- | --- | --- | --- |
|  | With<br>Heparin/LMWH | Without<br>Heparin/LMWH | Total | p-value* |
|  | (N=135) | (N=39) | (N=174) |  |
| **CD40 levels**<br>**(pg/mL)** |  |  |  | 0.8747 |
| N | 135 | 39 | 174 |  |
| Mean (SD) | 3836.9 (2266.12) | 3875.1<br>(2835.72) | 3845.4<br>(2396.70) |  |
| Median | 3194.3 | 2667.1 | 3138.7 |  |
| Range | 1234.3 - 14039.5 | 954.1 - 16706.1 | 954.1 -<br>16706.1 |  |
| **P-selectin**<br>**levels**<br>**(pg/mL)** |  |  |  | 0.7351 |
| N | 135 | 39 | 174 |  |
| Mean (SD) | 32996.4<br>(10181.13) | 34598.8<br>(9671.12) | 33355.6<br>(10063.86) |  |
| Median | 32622.1 | 35149.6 | 33593.0 |  |
| Range | 13979.9 -<br>73377.8 | 13586.4 -<br>59115.9 | 13586.4 -<br>73377.8 |  |

**Table 4:** sP-sel and sCD40L are not influenced by aspirin therapy after Aeson bioprosthetic total artificial heart (Aeson; A-TAH) implantation in 16 patients. *p-values from the linear mixed model

|  | Medication |  |  |  |
| --- | --- | --- | --- | --- |
|  | With Aspirin | Without Aspirin | Total | p-value* |
|  | (N=114) | (N=60) | (N=174) |  |
| CD40 levels (pg/mL) |  |  |  | 0.6823 |
| N | 114 | 60 | 174 |  |
| Mean (SD) | 3755.2<br>(2332.98) | 4016.9<br>(2524.51) | 3845.4<br>(2396.70) |  |
| Median | 3020.8 | 3410.2 | 3138.7 |  |
| Range | 1234.3 -<br>14039.5 | 954.1 -<br>16706.1 | 954.1 -<br>16706.1 |  |
| P-selectin levels (pg/mL) |  |  |  | 0.8839 |
| N | 114 | 60 | 174 |  |
| Mean (SD) | 33386.9<br>(9749.21) | 33295.9<br>(10720.66) | 33355.6<br>(10063.86) |  |
| Median | 33649.6 | 33161.8 | 33593.0 |  |
| Range | 13979.9 -<br>67263.4 | 13586.4 -<br>73377.8 | 13586.4 -<br>73377.8 |  |

**Figure 1:**
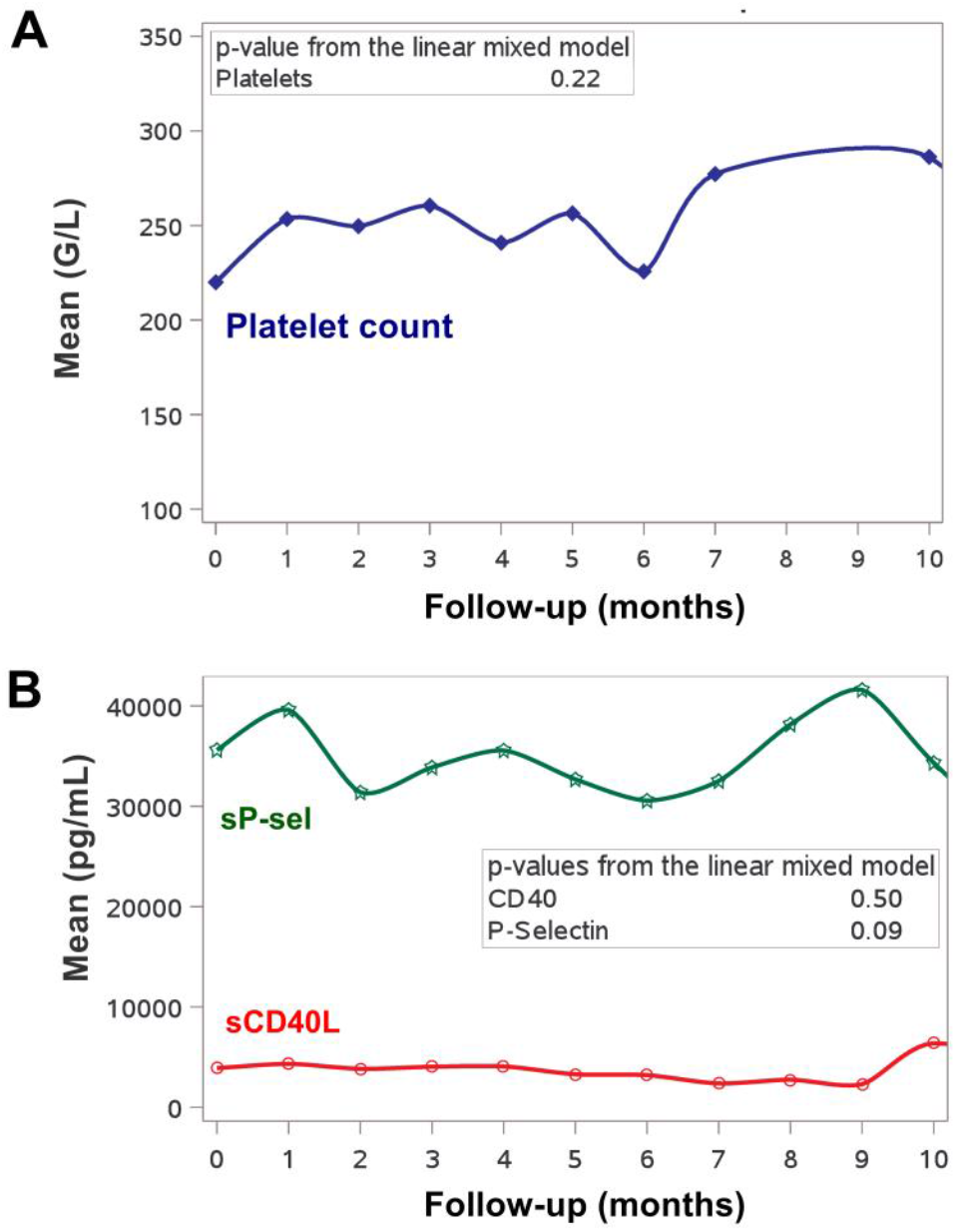
Absence of modification in platelet levels and platelet activation markers of activation after bioprosthetic total artificial heart (Aeson; A-TAH) implantation in 16 patients.

### Aspirin did not influence platelet activation biomarkers or pericardial effusion

Since platelet activation biomarkers have been related to high residual platelet reactivity during antiplatelet therapy ^14^, we analyzed levels of sP-sel and sCD40L according to aspirin use. As shown in Table 3, aspirin intake did not modify biomarker levels. No relation was found between pericardial effusion and aspirin use according in particular to aspirin start and drain removal (Table 5). The occurrence of pericardial effusion according to aspirin started before or after drain removal did not differ between the two groups (Fisher’s exact test, p-value=0.61). Moreover, the number of re-interventions for pericardial effusion according to aspirin start before or after drain removal did not either differ between groups (Mann-Whitney test, p-value=0.26).

**Table 5:** Link between AAP and pericardial effusion

| Patient | Aspirin start day after implantation | Drain removal day | Aspirin start before drain removal | Pericardial effusion | Number of re-interventions for pericardial effusion |
| --- | --- | --- | --- | --- | --- |
| 1 | Day 2 | Day 13 | Yes | No | 0 |
| 2 | Day 6 | Day 8 | Yes | Re-intervention at day 13 (940 ml) | 1 |
| 3 | Day 6 | Day 12 | Yes | No | 0 |
| 4 | Day 3 | Day 6 | Yes | Re-intervention at day 9 (530 ml) | 1 |
| 5 | Day 5 | Day 10 | Yes | No | 0 |
| 6 | Day 14 | Day 4 | No | No | 0 |
| 7 | Day 4 | Day 7 | Yes | 415 ml estimated on CT scan day 13; no intervention | 0 |
| 8 | Day 26 | Day 13 | No | No | 0 |
| 9 | Day 31 | Day 9 | No | No | 0 |
| 10 | Day 6 | Day 5 | No | 780 ml estimated on CT scan day 14 ; no re-intervention | 0 |
| 11 | Day 4 | Day 9 | Yes | Yes, re-intervention at day 14. | 1 |
| 12 | Day 7 | Day 7 | No | No | 0 |
| 13 | Day 8 | Day 4 | No | Yes, onset at day 29. Re-intervention at day 30. | 2 |
| 14 | Day 24 | Day 6 | No | Yes, onset at day 8. No re-intervention. | 0 |
| 15 | No aspirin | Day 8 | No | No | 0 |
| 16 | Day 10 | Day 5 | No | No | 0 |

## Discussion

In the present study, we provide additional proof of the high hemocompatibility of the pulsatile A-TAH device. We investigated the potential relationship between platelet activation, antithrombotic treatment, and outcomes after A-TAH implantation. We show that implantation of the A-TAH does not increase level of platelet activation biomarkers, in line with the safety of A-TAH implantation in terms of potential hemocompatibility-related events. Moreover, this level of activation marker is not influenced by the regimen of antithrombotic therapy used (aspirin or heparin/LMWH).

The lack of hemocompatibility has been largely attributed to shear stress inducing hemolysis, platelet activation and AWVS. Platelet activation has been largely described after LVAD implantation ^15–17^ and may result from interaction with artificial surface and flow related mechanisms. First, concerning artificial surfaces, LVADs are made of hemocompatible biomaterials such as titanium ^18^, which is relatively inert and prevents platelet adhesion. Titanium surfaces are not fully hemocompatible even with appropriate anti-thrombotic therapy since fibrinogen has been shown to be adsorbed onto the titanium ^19^. In contrast, larger surface in contact with blood in A-TAH is bovine-derived pericardial tissue which is widely used in bioprosthetic valve for the past 30 years ^20^. This surface can be covered by newly formed endothelial cells within a few weeks as previously described and probably confers anti-thrombotic properties to hybrid membrane of A-TAH ^21^. Second, platelet activation could be induced by flow related mechanisms. Indeed, shear-mediated platelet activation has been described as the result of a range of cell mechano-biological mechanisms whose mechano-activation is one of the main triggers. We previously described that 50 Pa is the threshold for platelet activation in mechanical circulatory support and corresponds to the threshold for approximately one second of exposure ^12^. In HMII and HM3 respectively 4% and 2.5% of the blood volume is concerned for shear stress over 50 Pa ^22^. In contrast we demonstrated that in the A-TAH these values are 100 times lower with less than 0.001% for left and 0.00015% for right ventricle regarding shear stresses >50Pa ^12^. Thus, the low shear stress could explain the absence of platelet activation after A-TAH implantation. This result is in line with absence of hemolysis and/or acquired von Willebrand syndrome during A-TAH ^13,21^.

Moreover, patients undergoing LVAD placement are at risk of pericardial effusion and subsequent cardiac tamponade because of the need for early postoperative antithrombotic treatment. After A-TAH, a tamponade could be a life-threatening condition because it could obstruct blood flow into the device and hence reduce cardiac output. Thus, absence of relation between pericardial effusion and aspirin is in line independence between clinical hemostatic events and platelet activation and/or an aspirin effect.

In MCS, thrombosis has been mainly attributed to platelet activation and hemolysis ^23^. Absence of platelet activation and hemolysis after A-TAH implantation is probably at least in part at the origin of safety in terms of hemocompatibility in implanted patients. In recent years, more than their involvement as main primary hemostatic actors, platelets have been extensively explored in inflammation and tissue repair and are now considered as bridge between hemostasis, inflammation, and innate immunity ^24^. Thus, platelet activation may lead and/or be associated to inflammation to promote thrombosis: this phenomenon so-called thrombo-inflammation leads to chronic inflammation. In patients supported with LVADs, chronic inflammation and immune dysfunction ^25–28^ associated with granulocyte activation ^29^ have been described. However, we previously described the absence of chronic inflammation after A-TAH implantation ^30^. Lack of platelet and chronic inflammation activation after A-TAH implantation result in a non-thrombotic environment which may offer an advantage over currently methods for biventricular support with LVADs implanted in both ventricles. Further, no data exist on biological markers of platelet activation after Syncardia implantation, however simulations with numerical models demonstrated increased stress-accumulations that could activate platelets ^31^ while simulations conducted on A-TAH observed absence shear stresses below thresholds of platelet activation ^12^.

When A-TAH clinical trials started, antithrombotic strategies similar to those for LVADs recommended antiplatelet agents (aspirin) and anticoagulation. The relative contribution of platelets and coagulation activation was unknown.

While anticoagulation at intermediate level seems mandatory ^3,5^, the need for antiplatelet agents in MCS and in particular after A-TAH is questionable. In LVAD, the question of antiplatelet treatment already exists since several European centers decided to decrease the dose or withdraw them ^6^. Indeed, TRACE and PREVENT II trials ^7,8^ proposed that deleting antiplatelet therapy is safe in terms of thrombotic events for HM2. The MOMENTUM 3 trial for HM3 demonstrated that low dose aspirin is also safe in terms of thrombotic events after HM3 implantation ^9^ suggesting aspirin withdrawal after HM3 implantation ^32^, which is now being tested in a dedicated trial ^33^. Since we demonstrated here that A-TAH is not associated with platelet activation, patients with A-TAH could likely benefit from reduced antithrombotic strategies using LMWH without aspirin, if their underlying disease do not require antiplatelet agents. LMWH continuation remains a real point of discussion because of the potential increased risk of bleeding in particular during transplantation. We have previously demonstrated that thrombocytopenia occurring after implantation is in general the result of disseminated intravascular coagulation due to interruption of anticoagulation. The timing of LMWH cessation is a matter of debate after A-TAH implantation ^34^. We have demonstrated here that LMWH/heparin do not influence level of platelet activation. The perfect MCS in terms of hemocompatibility will be the one without anticoagulation. The need of anticoagulation could be linked to existence of complete endothelialization of the hybrid membrane used in A-TAH. Prospective data in next clinical trials will evaluate percentage of newly formed endothelial cells on top of hybrid membrane to further understand if a complete stop of anticoagulation is possible.

All in all, results of this study demonstrate the safety of A-TAH implantation in terms of platelet activation. Absence of relationship between aspirin and platelet activation levels or pericardial effusion makes it attractive to consider circumventing antiplatelet therapy after A-TAH implantation. Additional clinical investigations are needed to test the value of aspirin treatment to further improve outcomes for long-term A-TAH patients.

## Data Availability

All data produced in the present study are available upon reasonable request to the authors

## Acknowledgment

We would like to thank the medical and nursing staff of cardiovascular intensive care units, cardiology and surgery departments involved in patient’s follow-up.

## Role and contribution of each author

DMS supervised the work, analyzed the data and wrote the paper. CP, LJ and AP performed analysis and reviewed the paper. PI, FZR, YP, CL, FG, JCR, AV, EF, PG and IN included patients, analyzed the data and reviewed the paper. MG, CL and PJ provided data and reviewed the paper.

